# The effect of proactive, interactive, two-way texting on 12-month retention in antiretroviral therapy: findings from a quasi-experimental study in Lilongwe, Malawi

**DOI:** 10.1101/2024.01.26.24301855

**Authors:** Caryl Feldacker, Robin E. Klabbers, Jacqueline Huwa, Christine Kiruthu-Kamamia, Agness Thawani, Petros Tembo, Joseph Chintedza, Geldert Chiwaya, Aubrey Kudzala, Pachawo Bisani, Dumisani Ndhlovu, Johnnie Seyani, Hannock Tweya

## Abstract

**Background:** Retaining clients on antiretroviral therapy (ART) is challenging especially during the first year on ART. Mobile health (mHealth) interventions show promise to close retention gaps. We aimed to assess reach (who received the intervention?) and effectiveness (did it work?) of a hybrid two-way texting (2wT) intervention to improve ART retention at a large public clinic in Lilongwe, Malawi.

**Methods:** Between August 2021 - June 2023, a quasi-experimental study compared outcomes between two cohorts of new ART clients: 1) those opting into 2wT with combined automated, weekly motivation short messaging service (SMS) messages and response-requested appointment reminders; and 2) a matched historical cohort receiving standard of care (SoC). *Reach* was defined as “the proportion clients ≤6 months of ART initiation eligible for 2wT”. 2wT *effectiveness* was assessed in time-to-event analysis comparing Kaplan-Meier plots of 6- and 12-month retention between 2wT and SoC using a log-rank test. The effect of 2wT on ART drop out was estimated using multivariable Cox proportional hazard models, adjusting for sex, age and WHO stage at ART initiation.

**Results:** Of the 1,146 clients screened, 645 were ineligible (56%) largely due to lack of phone access (393/645; 61%) and illiteracy (149/645; 23%): a reach of 44%. Among 468 2wT participants, the 12-month probability of ART retention was 91% (95%CI: 88% - 93%) compared to 75% (95%CI: 71% - 79%) among 468 SoC participants (p<0.0001). Compared to SoC participants, 2wT participants had a 62% lower hazard of dropping out of ART care at all time points (hazard ratio 0.38, 95% CI: 0.26-0.54; p<0.001).

**Conclusions:** Not all clients were reached with 2wT. For those who opted-in, 2wT reduced drop out throughout the first year on ART and significantly increased 12-month retention. The proactive 2wT approach should be expanded as a complement to other interventions in routine, low-resource settings to improve ART retention.

## Introduction

In sub-Saharan Africa (SSA), persistent gaps in retention on antiretroviral therapy (ART) among specific groups and geographies of people living with HIV (PLHIV) threaten decades of impressive progress [1]. Loss to follow up (LTFU) is highest among those newly on ART through 6 months in care [2, 3]. Gaps in retention are a problem: even short treatment interruptions can lead to increased client morbidity, mortality, drug resistance, and HIV transmission risk [4]. Most efforts to address LTFU are reactive, waiting for clients to miss visits before intervening, resulting in tracing delays that reduce the likelihood of finding, returning, and retaining clients in care [5, 6]. Addressing retention gaps is costly, often relying on healthcare workers (HCWs) to call or trace clients identified as LTFU in-person [7]. Recent reductions in global funding, chronic shortages of HCWs, and increasing client volumes exacerbate retention challenges. Proactive, lower-intensity, effective retention interventions, especially for ART clients in their first year in care, are needed in routine, low-resource SSA settings.

Numerous mobile health (mHealth) interventions show promise to significantly increase ART retention (alive in care, adherence to ART, visit compliance) among adults [8-14]. Effectiveness, however, is not guaranteed; not all mHealth interventions are associated with improved ART retention [15-19]. Previous mHealth intervention research suggests that several mHealth intervention characteristics raise the likelihood of ART retention impact. First, lower technology approaches that rely on short-message service (SMS) which requires only feature phones appear better suited to low- or middle-income country (LMIC) settings [20, 21]. SMS-focused interventions show high acceptability [11, 22-24] and reduce digital health equity concerns associated with apps that require smartphones [25]. Second, interactive interventions that enhance communication between clients and HCWs are more effective than one-way blast communication to engage clients in care [13, 26]. Interaction potentially diminishes message fatigue and facilitates more personalized, intensive support when needed [27]. Third, engagement of diverse stakeholders throughout the design, testing, and evaluation process creates ownership and buy-in [28], helping tailor the right interventions to the local context [29]. Lastly, iterative monitoring and evaluation (M&E) of mHealth in accordance with digital health best practices suggested by the World Health Organization (WHO) helps to ensure these interventions complement, as opposed to conflict with, ongoing health system strengthening [30, 31].

Malawi is an ideal location to assess mHealth to improve client retention. Malawi is a low-resource country with an adult HIV prevalence of ∼7% [32]. Although progress towards 95-95-95 appears on track [32], LTFU is high, especially during in the first year on ART [33]. Five years after ART initiation, only 54% of PLHIV are retained in ART care [34]. Given pervasive HCW shortages [35], alternatives to human resource-intensive solutions are needed. In Malawi, the Ministry of Health (MoH) has tested several ART-related innovations at Lighthouse Trust (LT), one of the largest ART providers and a WHO-recognized ART Centre of Excellence in Lilongwe, the capital [36]. LT provides integrated HIV care to 38,000 ART clients in its two flagship clinics in urban Lilongwe: 25,000 at Martin Preuss Centre (MPC) and 13,000 at Lighthouse clinic (LH). At LT, retention at twelve months post-ART initiation is estimated at 73%, falling to 63% by 24 months. In 2006, LT established an intensive client retention program, “Back-To-Care” (B2C), in which up to three calls or home visits are attempted to encourage clients who miss their scheduled appointment to return to care; B2C currently aims to trace all clients who miss visits by ≥14 days. B2C has been well-recognized for its success returning clients to care [37-41]. However, B2C, like other reactive tracing efforts, is highly resource intensive [38]. At MPC clinic from July-September, 2023, there were 18,842 scheduled ART visits; 1,798 clients (10%) missed visits by ≥14 days and were referred to B2C. With five, full-time B2C tracers, only 40% (719/1798) of potential LTFU clients during that period were successfully found. B2C efforts are stretched, leading to delayed or missed tracing. LT needs proactive, effective, retention innovations that reflect the reality of routine, low-resource, public settings.

In 2021, LT and partners at the University of Washington’s International Training and Education Center for Health (I-TECH) and Medic developed a two-way texting (2wT) system to improve early retention at MPC clinic. 2wT is an hybrid (automated and interactive) intervention combining weekly non-HIV-related motivational messaging and response requested scheduled ART visit reminders, aiming to provide proactive retention support to prevent care gaps before they happen. Early 2wT usability assessment among new ART initiates (within 6 months of initiation) demonstrated high client acceptability and support for the 2wT approach [42].

We aimed to assess the impact of the 2wT intervention on 12-month retention among new ART initiates at MPC using a quasi-experimental design. We employed an implementation science (IS) approach to enhance the quality, speed, and impact of translating 2wT research findings into routine practice [43]. We applied the RE-AIM framework (reach, effectiveness, adoption, implementation, maintenance) to guide our evaluation[44] and further understanding for whom, where, why and how 2wT works [45]. In this paper, we assess 2wT *reach* by describing the participant flow from 2wT screening, eligibility, and enrollment and examine 2wT *effectiveness* by comparing ART retention at 12 months between 1) new ART clients who opted into 2wT (intervention) versus 2) a historical cohort of routine MPC new initiates who received standard of care (SoC) (comparison). We hypothesized that 2wT would improve 12-month retention by ≥10%, from 73% to 83%.

## Methods

### Setting

The study was conducted at MPC, LT’s largest urban clinic in Lilongwe, Malawi. LT operates as part of MoH ART service delivery and employs the MoH electronic medical record system (EMRS) at MPC [46]. On average, MPC initiates 450 PLHIV on ART per quarter following the test and treat strategy. At ART registration, clients’ demographics, phone number(s) and WHO stage are captured in the EMRS [47]. During the first three to six months after ART initiation, clients are seen monthly, after which, if clients are stable and adherent to ART, visit frequency is typically decreased to once every three- or six-months.

### Study design, cohort creation and sample size

A quasi-experimental design was used to assess the effect of 2wT on 12-month ART retention by comparing retention among a cohort of 2wT participants to that among a matched historical comparison cohort receiving SoC at MCP one year prior to 2wT implementation. The intervention cohort was matched 1:1 on age (bands of 5 years), sex, and WHO stage at initiation via random selection from an MPC dataset of 1,455 ART clients who had a phone number. With a baseline 12-month retention at MPC of 73% in March 2021, 438 participants would be required in each arm to provide 90% power to detect a 10% retention difference between 2wT and SoC. We recruited 501 2wT participants to account for transfer-out, withdraws and deaths. 2wT participants-initiated ART between May 2021 – April 2022 and were followed through 12 months post-ART initiation. The historical, matched, comparison SoC cohort initiated ART at MPC between November 2019 - November 2020, before 2wT launch.

### 2wT behavior change theory

2wT design was informed by the theory that prompts can spur action to change [48]. By using SMS to target key individual-level constructs proven effective in previous HIV-related behavior change programs [49-52] it is expected that the 2wT intervention will improve early retention on ART (**Fig 1**). To increase *behavioral control* over timely attendance at scheduled ART visits, participants are reminded by SMS in advance to provide time to arrange transport or free their schedules. 2wT helps improve participant *motivation* to make decisions for their own wellness, including adhering to ART, via weekly non-HIV (neutral) messages or educational content that support participant engagement in their health. Initial 2wT participant education and subsequent interaction with 2wT officers encourages *self*-*efficacy* by providing participants with an opportunity to request visit date changes, report transfers, or communicate about any issues related to their visits.

**Fig 1:**
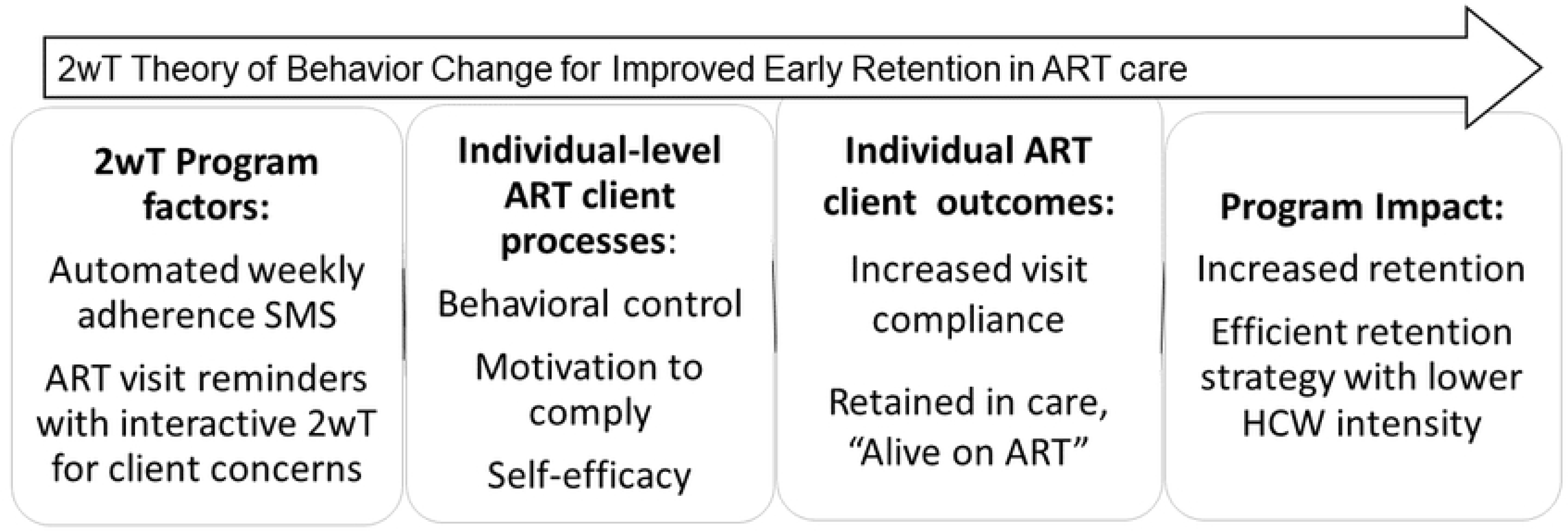
Program theory of change for 2wT.

## Retention interventions

### Standard of care (SoC)

The historical comparison cohort received SoC at MPC. As of mid-2019, as part of SoC, all new ART initiates at MPC were assigned an *ART Buddy*, a PLHIV on ART at LT who would serve as a “buddy” for their first 12 months on ART. Buddies are considered Expert Clients, a cadre of paid, trained, peer-supporters who guide clients through early ART care by providing health education, disclosure support, and adherence counseling. Each ART Buddy is paid a nominal fee to support ∼15 new ART initiates.

The 2wT and historical comparison cohort received different retention support (**Table 1**).

**Table 1:** Retention support received by 2wT and SoC participants.

| Activity | First 12 months on ART |  |
| --- | --- | --- |
|  | Standard of Care (SoC) | Two-way texting (2wT) |
| **At ART Initiation** |  |  |
| ART education and counselling at ART initiation | Yes | Yes |
| Completion of locator form for B2C purposes | Yes | Yes |
| Assigned an ART Buddy | Yes | No |
| **Proactive retention activities** |  |  |
| Weekly motivational messages (general encouragement and information about healthy habits) | No | Yes |
| Before a visit | Phone calls by an ART Buddy 3 days and, if necessary, 2 days and 1 day before a scheduled visit | Automated SMS visit reminders 3 days and 1 day before a scheduled visit |
| **Reactive retention activities** |  |  |
| 1-13 days after a missed visit (reactive) unless a client returns to care | Up to 3 phone call attempts by ART Buddy after a missed visit | Automated SMS missed appointment reminders on days 2, 5, and 11 after scheduled visit |
| Back to care (B2C) referral for tracing $\geq 14$ days | Weekly manual generation of tracing list | Daily automated generation of tracing list |

### 2wT intervention

The 2wT approach was developed in accordance with the Principles for Digital Development [53] and based on the open-source Community Health Toolkit (CHT) [54]. 2wT’s easy-to-use CHT-based design resulted from an iterative human-centered design (HCD) process that incorporated feedback from LT clients, HCWs, and retention officers, described previously [55]. In brief, 2wT is a free, proactive, mHealth intervention that combines automated motivation messages with interactive individualized visit reminders (**Fig 2)** [42]. 2wT does not use an app, nor does it require participants to download anything. ART clients who opt into 2wT are sent weekly one-way, “blast” motivation messages containing non-HIV-related content such as generic messages of encouragement (e.g., “You can do it”, “You are making great choices every day”) and general health advice (e.g., “Drink boiled and clean water”, “Seek help when you do not feel well”). Additionally, 2wT participants receive response-requested individualized visit reminders day 3 and 1 before their scheduled clinic visit. Participants who confirm visit reminders with a “yes” end the dialogue. A “no” triggers interactive SMS with a HCW in which participants may change their appointment dates, report transfers, or chat about other visit-related issues. Participants who miss a scheduled clinic visit are sent follow-up reminders 2, 5, and 11 days after the appointment (unless they return to care). Messages are stopped upon participant request, transfer or death.

**Fig 2:**
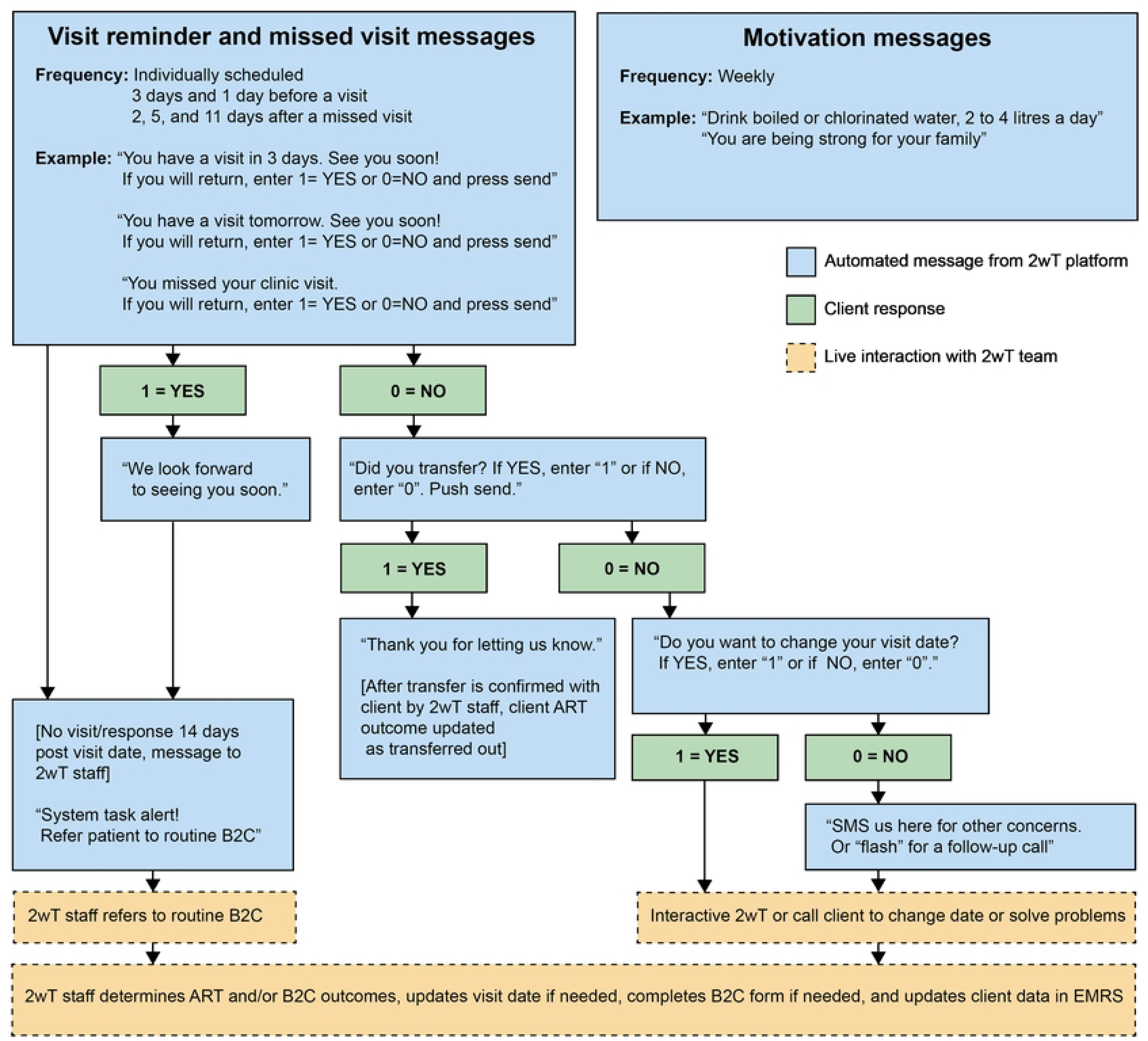
2wT intervention message flow.

### 2wT recruitment: screening and enrollment

During the 2wT enrollment period (August 2021 - April 2022), all new ART clients at MPC were screened for study eligibility, including: 1) initiated ART <6 months prior; 2) ≥18 years; 3) possessed phone at enrolment; 4) had basic literacy; 5) completed informed consent; and 6) received their 2wT enrollment text. Eligible participants enrolled to receive messages in either Chichewa or English, based on their preference, but could send SMS in any language. Participants received instructions on how to respond to 2wT messages.

## Data collection

For both the 2wT and SoC cohort, data on routine MoH ART outcomes were extracted from the EMRS for participants’ first twelve months post-ART initiation (May 2021 – June 2023 for 2wT and November 2019 – January 2022 for SoC). These outcomes could be either 1) *alive and on ART* (alive and retained in ART care on the date of record review); 2) *stopped ART treatment* (alive but informed the clinic they stopped ART); 3) *transferred* (documented move to another facility); 4) *dead* (all cause mortality); and 5) *LTFU* (no return to clinic within 60 days of a scheduled visit) [46]. To ensure correct ART outcome ascertainment, intervention and comparison group outcomes were updated for 60 days beyond the 12-month period. Participants who requested to stop visit reminder messages were considered to have withdrawn from the study and were classified as *withdrew*. SMS data was obtained from the 2wT database and the SMS aggregator, Africa’s Talking.

## Study Outcomes

*Reach* was measured using screening data and was defined as the proportion of screened PLHIV eligible and willing to participate in 2wT. *Effectiveness* was measured as the effect of 2wT on 12-month retention on ART, defined as “alive on ART” according to the Malawi MoH ART outcome, as opposed to “drop out”, a study-specific grouping that included both “stopped ART treatment” and “LTFU”. We examined message response rates and 6-month retention on ART care as secondary analyses.

## Statistical analysis

2wT participants were considered exposed to the intervention if at least one visit reminder SMS was successfully delivered to their phone within the 12-month follow-up period; exposed participants were included in analysis. Descriptive statistics present matching success between the 2wT and SoC cohort, MoH ART outcomes at 6- and 12-month follow-up for 2wT vs. SoC clients, and describe 2wT reach. Counts and frequencies report the number of messages sent by the 2wT platform and by participants as well as message response rates. Chi-square tests were performed to compare the proportion of participants in each MoH ART outcome category for the 2wT and SoC cohort. 2wT participants who withdrew from the study were excluded from the denominator. Effectiveness was primarily assessed in time-to-event analysis. Transfer, death and withdrew were censoring events; LTFU and stopping ART treatment were failure events. Participants were censored twelve months after ART initiation. Individuals in the intervention arm who enrolled in the study after ART initiation were considered to come under observation at the time of enrolment but entered analysis taking account of the accumulated time on ART. Kaplan-Meier plots of retention on ART and probabilities of being retained on ART six and twelve months post-ART initiation are presented, comparing 2wT and SoC retention curves using a log-rank test. The effect of the intervention on ART drop out was estimated using Cox proportional hazard models. Multivariable Cox proportional hazard models were used to model the association between retention intervention (2wT or SoC) and ART drop out, adjusting for sex, age and WHO stage at ART initiation. We assessed the proportional hazards assumption using Schoenfeld residuals and assessed interaction between the intervention and relevant covariates. The unadjusted hazard ratio (HR) and adjusted HR were reported with 95% confidence interval.

## Ethics

The study protocol was approved by the Malawi National Health Sciences Research Committee (#20/06/2565) and the University of Washington, Seattle, USA ethics review board (STUDY000101060). At enrolment, 2wT participants provided written informed consent in either Chichewa or English, according to their preference. SoC clients did not consent as only de-identified, routine monitoring and evaluation data was collected from EMRS.

## Results

### 2wT Reach

The study team screened 1,146 new ART clients at MPC to yield a cohort of 501 2wT participants, an intervention reach of 44% (**Fig 3**). The most common reasons for ineligibility were lack of phone access (393, 61%), illiteracy (149, 23%), and age under 18 years (48, 7%). The majority of 2wT participants were female (56%), had an average age of 33 years, and WHO clinical stages 1 or 2. Among 2wT participants exposed and included in analysis, 373 (80%) were enrolled at the ART initiation visit while the remaining 95 (20%) were enrolled during a subsequent visit, on average 99 days (standard deviation (SD): 53 days) after ART initiation. The matching process successfully created similarity in matched demographic and clinical characteristics between 2wT and SoC groups (**Table 2**).

**Fig 3:**
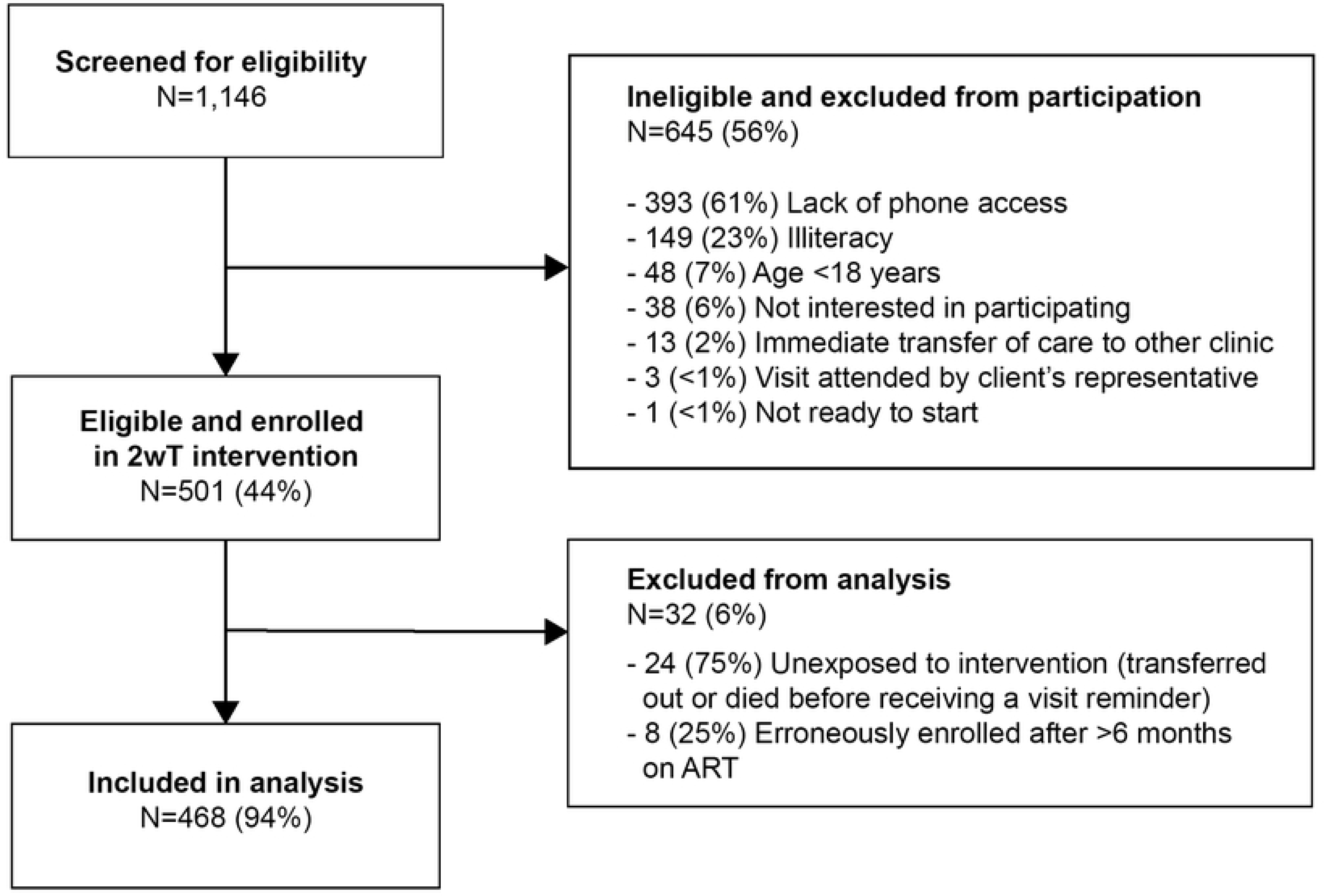
2wT enrollment flow: Screening, eligibility, and enrollment participation.

**Table 2.** Participant characteristics.

|  | 2wT arm<br>(N=468) | Standard of care<br>(N=468) |
| --- | --- | --- |
| Sex |  |  |
| Female | 261 (55.8%) | 261 (55.8%) |
| Male | 207 (44.2%) | 207 (44.2%) |
| Age (years) |  |  |
| Median [interquartile range (IQR)] | 33 [27 - 40] | 33 [27 - 40] |
| Age group (years) |  |  |
| <20 | 4 (0.9%) | 4 (0.9%) |
| 20-24 | 64 (13.7%) | 64 (13.7%) |
| 25-29 | 102 (21.8%) | 102 (21.8%) |
| 30-34 | 88 (18.8%) | 88 (18.8%) |
| 35-39 | 90 (19.2%) | 90 (19.2%) |
| 40-44 | 56 (12.0%) | 57 (12.2%) |
| 45-49 | 42 (9.0%) | 41 (8.8%) |
| 50-54 | 11 (2.4%) | 11 (2.4%) |
| 55-59 | 7 (1.5%) | 7 (1.5%) |
| 60-64 | 2 (0.4%) | 2 (0.4%) |
| 65-70 | 1 (0.2%) | 2 (0.4%) |
| 75+ | 1 (0.2%) | 0 (0%) |
| WHO stage |  |  |
| Stage 1 or 2 | 364 (77.8%) | 364 (77.8%) |
| Stage 3 | 71 (15.2%) | 71 (15.2%) |
| Stage 4 | 30 (6.4%) | 30 (6.4%) |
| Unknown | 3 (0.6%) | 3 (0.6%) |
| Time on ART before enrollment |  |  |
| Median [Min, Max] | 0 [0, 182] | 0 [0, 29.0] |

### 2wT platform engagement

During the study period, the 2wT platform recorded a total of 31,861 messages. The 2wT system sent 27,859 SMS (87%): 18,093 motivational messages (65%); 4,561 visit reminders (16%); 1,468 missed visit reminders (5%); and 3,737 other messages (13%) (**Fig 4**). The delivery success rate was 76% for motivation messages; 79% for visit reminders; and 75% for missed appointment reminders. Of all 31,862 messages, 4,002 messages (13%) were sent by participants. Participants responded to 39% of successfully delivered pre-visit reminders (proactive) and 32% of successfully delivered post-missed visit reminders (reactive). Of 16 (3%) participants who requested to stop 2wT messaging, 5 lost interest (31%), 5 noted confidentiality concerns (31%), and 3 noted no longer needing texts to remember their appointments (19%) as reasons to stop.

**Fig 4:**
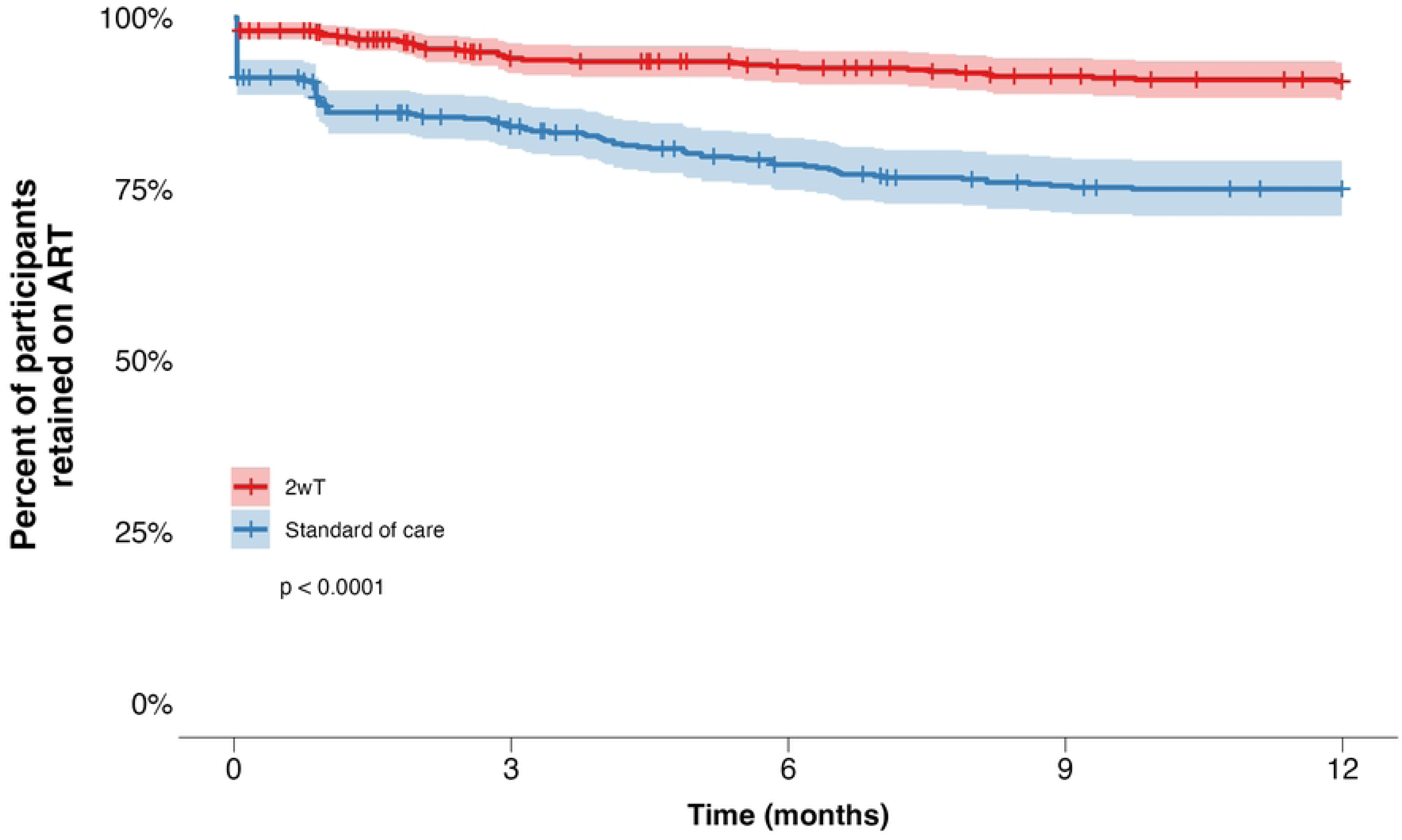
Interactive SMS flow between the 2wT system and the participants.

### 2wT effectiveness

#### ART outcomes at 6 and 12 months

Six months post-ART initiation, 88% were alive and in care in the 2wT arm as compared to 76% in the SoC arm (p<0.001). The 2wT arm had lower LTFU (5% vs. 11%) than SoC (p<0.001) and lower stopped treatment (1% vs. 5%) (p<0.001) than SoC arm (Table 3). At 12 months post-ART initiation, 81% of 2wT were alive and in care as compared to 66% of SoC (<0.001). The 2wT arm had lower LTFU (7% vs 18%) (p<0.001) and stopped treatment (2% vs 6%) (p=0.004) compared to SoC. Equal percentages of both 2wT and SoC transferred out (9% vs 9%) and died (2% vs 2%) at twelve months.

**Table 3:** 6- and 12-month MoH ART outcomes of 2wT vs. SoC participants.

|  | **2wT arm**<br>**N = 468** | **Routine**<br>**N = 468** |  |
| --- | --- | --- | --- |
|  | N (%) | N (%) | P-value |
| 6-months post-ART initiation | 454* | 468 |  |
| Alive | 401 (88) | 354 (76) | <0.001 |
| LTFU | 21 (5) | 52 (11) | <0.001 |
| Stopped treatment | 6 (1) | 25 (5) | <0.001 |
| Transfer out | 21 (5) | 30 (6) | 0.2 |
| Dead | 5 (1) | 7 (2) | 0.6 |
| 12-months post ART-initiation | 452* | 468 |  |
| Alive | 365 (81) | 309 (66) | <0.001 |
| LTFU | 31 (7) | 84 (18) | <0.001 |
| Stopped treatment | 10 (2) | 28 (6) | 0.004 |
| Transfer out | 39 (9) | 40 (9) | 1 |
| Dead | 7 (2) | 7 (2) | 0.9 |

\*By 6-months, 14 participants, and by 12-months, 16 participants had withdrawn from the study

#### Retention in care at 6 and 12 months

Kaplan-Meier curves revealed differences in retention on ART over time between the 2wT and the SoC arm (p<0.0001) (**Fig 5**). Six months post-ART initiation, the probability of being retained on ART in the 2wT group was 93% (95% CI: 91% - 95%) compared to 79% (95%CI: 75% - 82%) in the SoC group. At twelve months, the probability of retention on ART was 91% among 2wT participants (95% CI: 88% - 93%) and 75% among SoC participants (95% CI: 71% - 79%).

**Fig 5:**
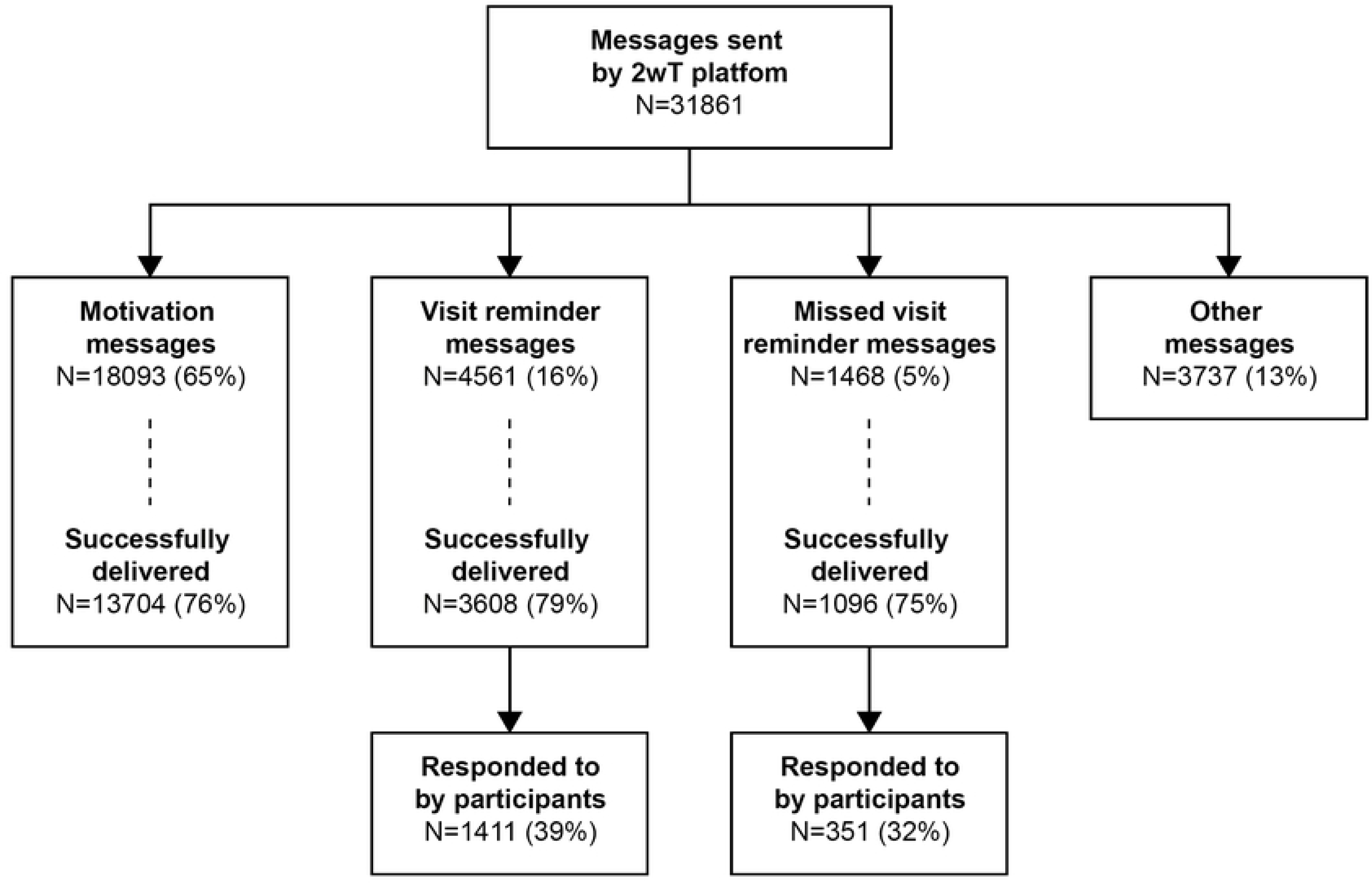
Kaplan-Meier curve of retention on ART among 2wT and SoC clients over time, displaying 95% confidence intervals and log-rank test p-value.

#### Drop out during the first year on ART

In unadjusted analysis, being in 2wT arm was associated with a 62% lower hazard of drop out during follow-up (HR 0.38, 95%CI: 0.27 – 0.55) as compared to SoC. A similar effect on drop out was found controlling for sex, age and WHO stage (HR 0.38, 95% CI: 0.26-0.54). In the first year on ART, men were more likely to drop out of care: in the multivariate analysis, being male was associated with a 49% higher hazard of dropout from ART care as compared to females (HR 1.49, 95% CI 1.07-2.09). Each additional year of age was associated with a 5% reduction in hazard of dropout (HR 0.95% CI:0.93-0.97). Compared to individuals with WHO stage 1 or 2, those with WHO stage 3 at ART initiation had a 5% lower hazard of dropout (HR, 0.95% CI 0.59-1.52) while those with WHO stage 4 had a 31% higher hazard (HR 1.31, 95% CI 1.07-2.09).

## Discussion

As a result of the combined motivation messages and automated visit reminders of the 2wT intervention, retention in ART care at 12 months was 16% higher among 2wT participants as compared to SoC clients. At any point over the first year on ART, those with 2wT support were 62% less likely to drop out as compared to their SoC peers. For both 2wT and SoC, men were more likely to drop out than females, and older clients were more likely to be retained. Improved retention among 2wT participants is most striking in the first months immediately after ART initiation, a high risk period when care gaps are most likely [56]. Still, 2wT effectiveness should not eclipse limitations in 2wT reach: over 50% of those invited to participate were ineligible, largely due to lack of phone access or illiteracy. This evidence informs discussion of the strengths and weaknesses of the 2wT approach and provides guidance for transitioning the optimization of 2wT from research to routine practice.

Several aspects of 2wT design likely contributed to success. First, we used a human-centered, co-design process with clients, HCWs, and stakeholders, informing iterative improvements that likely raised the likelihood of matching the right 2wT design to the expressed local need [57]. For participants, the 2wT approach was perceived as user-friendly, responses were easy, and both motivational and visit reminders were appreciated [42]. For HCWs, 2wT was highly usable and was perceived to improve their connection with participants, reflecting their needs [55]. Lighthouse HCWs currently maintain the day-to-day operations and management of the 2wT system independently, factors that enhance the likelihood of sustaining the intervention [58]. The open-source 2wT technology, itself, appears to be the right fit for the low-resource setting. 2wT requires only basic phones with SMS capability from participants (no smartphones, no need to download apps, no data plan) and a web-based interface for HCWs that runs on commonly available PC computers and Android tablets. The workload also meets the low resource setting. Currently, one 2wT officer handles all client interaction for over 400 participants as compared to SoC where each ART Buddy is assigned up to 15 new ART clients. The hybrid automated and manual design relies heavily on 2wT automated reminders, both before visits and after missed appointments, requiring realistic effort while the intensity of direct participant-to-HCW interaction appears manageable.

The intervention messages and their scheduling also likely improved outcomes. First, 2wT messages were informed by health behavior theory, adapting the messages alongside software optimization to increase the strength of both [59]. Over two decades of research demonstrates the importance of having a strong theoretical model to provide rationale for how interventions may influence behaviors [60]. We suggest that 2wT content helped improve participant motivation, behavioral control, and self-efficacy, hoping to create positive habits of ART adherence from initiation onward. Second, the cadence of 2wT messaging intensity appears to suit participants. Although other mHealth with weekly response-requested messaging found no effect on 12-month retention [61], 2wT included weekly motivational or education messages without requesting a response and only required participants to interact with the system or HCWs with a single “1=yes” or 0=no” to confirm visits a few days before appointments, with option to interact more if needed or desired. The 2wT nudge and minimal participant effort likely lessened the potential fatigue that more messages could cause [13]. Furthermore, a previous mHealth qualitative study noted that fears of unintended disclosure from HIV-related message content could reduce SMS intervention participation or uptake [11]. Using suggestions from current 2wT users, 2wT may have found the right type of mixed educational and motivational content, without HIV-related messaging.

Despite effectiveness, effort will be needed to expand 2wT reach as only 44% of those screened for participation met the 2wT eligibility criteria. Current access in Malawi to mobile phones in 2022 was estimated at 60% [62] with access among females likely lower [63]. However, growth in mobile phone ownership is expected to rise, potentially reducing these concerns in future. Likewise, as more than 25% of adults in Malawi are unable to read or write [64], future voice functionality in 2wT or improved “flash” features (calling a number to trigger a voice call back) could expand reach in response to low literacy client needs [13]. As 2wT participants noted a preference for SMS over calls, given that messages are discrete and do not require a participant to pick up or attend to them at a specific time [42], voice should not replace SMS, but augment existing options. Additive retention models, where clients can have more than one retention support may also lead to gains [12], improving reach and impact as 2wT moves from research to routine practice. Expanding enrolment into 2wT for any client on ART, and including clients ages 15 and older (the age of consent in Malawi) combined with efforts to improve 2wT awareness among LT clients who come on evenings and weekends (when study team were not available) could improve 2wT uptake of 2wT retention support.

## Limitations

Our findings should be considered in light of limitations. First, it is possible that 2wT effect could diminish over time [65], and future studies of 2wT reach adaptation, flexibility, or optimization should explore how to maintain the early retention gains. Second, despite successful matching, using a historical comparison group may pose a threat to internal validity, especially given the different temporal influences of COVID-19 on participants. Third, the extent to which the two study cohorts received the intended retention support, i.e., whether 2wT participants actually received 2wT (were SMS read?) or whether PLHIV buddies actually called SoC participants (did ART Buddies provide intended support?) is unknown, calling for future fidelity investigation of both 2wT and SoC interventions in practice. Moreover, 2wT was opt-in, allowing those who were eligible and interested to volunteer for the retention support. Participants who choose an intervention are likely more open and responsive to the support. Lastly, we did not include the outcomes of those who transferred out as we lacked resources to track clients to other clinics. Despite these limitations, the strengths of this quasi-experimental design in the routine Malawi ART setting suggests that this specific 2wT approach may be beneficial to improve early retention among the sizeable population who wish to opt-in.

## Conclusions

At a high-volume, routine ART clinic in Malawi, the proactive, low-intensity 2wT approach improved 12-month retention among new ART initiates who enrolled. 2wT should be scaled as a part of, and not a replacement for, complementary retention efforts in routine ART settings in Malawi. Even with sub-optimal reach, adoption the 2wT approach as a component of routine retention efforts could benefit the ART client population as a whole by freeing existing HCWs to trace more clients presumed LTFU, returning more clients to care. More retention choices could also help cater to the diverse preferences and practicalities of retaining clients on ART over time. Given the large client volume of LT clinics, Lighthouse’s leadership, and continued MoH collaboration, expansion of the 2wT retention approach for both new and existing ART clients could positively impact overall ART program success at scale.

## Data Availability

All relevant data are within the manuscript and its Supporting Information files.

## Acknowledgements

We would like to acknowledge the study participants, the study team (Kondwani Masiye, Harrison Chirwa, Blessings Wandira, Daniel Mwakanema, Madalitso Chawanje, William Maziya, Isaac Nyirenda), colleagues from Medic (Maryanne Mureithi, Femi Oni, Beatrice Wasunna, Edwin Kagereki, Adinan Alhassan, Kawere Wagaba, Evelyn Waweru, and Mourice Barasa), the MPC clinic, M&E teams, B2C team, and MoH staff at Bwaila for their invaluable contribution to the study.

## Funding

Research reported in this publication was supported by the Fogarty International Center of the National Institutes of Health under Award Number R21TW011658/R33TW011658. The content is solely the responsibility of the authors and does not necessarily represent the official views of the National Institutes of Health.

## Data Availability statement

The data that support the findings of this study are included in supplemental materials.

## Supplemental information

**S1 File. Dataset.** Retention outcomes dataset in CSV format.

## Notes

### Competing Interest Statement

The authors have declared no competing interest.

### Author Declarations

Ethics The study protocol was approved by the Malawi National Health Sciences Research Committee (#20/06/2565) and the University of Washington, Seattle, USA ethics review board (STUDY000101060). At enrolment, clients provided written informed consent in either Chichewa or English, according to their preference.

